# HLA-DQB1*03:01 strongly affects age of onset of type 1 narcolepsy independently of DQA1 and ethnicity

**DOI:** 10.1101/2025.06.14.25328971

**Authors:** Lisan Zhang, Shuo Cai, Ashton Teng, Ling Lin, Fang Han, Han Yan, Seung-Chul Hong, Fabio Pizza, Giuseppe Plazzi, Ambra Stefani, Birgit Högl, Michel Lecendreux, Patrice Bourgin, Isabelle Arnulf, Stine Knudsen-Heier, Takashi Kanbayashi, Yu-Shu Huang, Poul Jennum, Karel Sonka, Sona Nevsimalova, Makoto Honda, Fernando Morgadinho Coelho, Zerrin Pelin, Luigi Ferini-Strambi, Taku Miyagawa, Seik-Soon Khor, Katsushi Tokunaga, Wanlu Liu, Emmanuel J Mignot

**Author notes:** Corresponding author: Emmanuel J Mignot, Stanford University, Center for Sleep Sciences and Medicine, Department of Psychiatry and Behavioral Sciences, Palo Alto, CA, 94304, USA. **Author Contributions:** Study design, L.Z, and E.-J.M. Sample collection, L.Z, L.L., F.H., H.Y., S.-C.H., F.P., G.P., A.S., B.H., M.L., P.B., I.A., S.K-H., T.K., Y.-S.H., P.J., K.S., S.N., M.H., F.-M.C., Z.P., L.F-S., T.M., S.-S.K., K.T., and E.M. Data generation, processing, and analysis, L.Z., L.L., S.C., A.T., W.L., and E.M. Manuscript preparation, L.Z., and E.M. Supervision, E.M. **Competing Interest Statement:** The authors declare that they have no competing interests.

## Abstract

In this study, we investigated the effects of HLA-DQB1*03:01 on the onset age of type-1 narcolepsy. We pooled data from 5,339 cases from China, Europe, Korea, Japan, and the United States to conduct the first transethnic study on age of onset (OA). GWAS was used to detect the associated genes. HLA alleles were imputed by HIBAG. Three datasets (DGN, CHINA, and CAUC) were used for TCR analyses. Chinese patients were found to have an earlier age of onset. Independently of this, only one strong GWAS significant effect was observed, summarized by the presence of DQB1*03:01 and centered around the coding region of this gene. In contrast, DQ0602 dosage did not strongly affect onset, and other known narcolepsy-associated genetic loci had minor effects. The DQB1*03:01 effect (mean −3.47 years, p=1.7 10-18) showed no heterogeneity across ethnic groups and was independent of common allelic variation at DQA1 in cis of DQB1*03:01 (DQA1*03:03; DQA1*0505; DQA1*06:01). This effect may be explained by presentation of peptide(s) across all DQ0301 heterodimers, or, considering genetic effects mapping on TCRA and TCRB, effects on TCR repertoire. Using bulk and single-cell RNA-seq data across two ethnic groups, we found that DQB1*03:01 alters TCR repertoire at specific positions, most significantly within the CDR2A, CDR2B and CDR3B loops. These results illustrate the remarkable conservation of narcolepsy genetic effects across ethnicity and suggest that specific TCRs regulate narcolepsy onset. This result will likely be illuminating of the pathophysiology once T cells causative of the disease have been identified.

## Introduction

Type 1 narcolepsy is characterized by sleepiness, cataplexy, weight gain and rapid-eye-movement sleep abnormalities. It affects ∼1/3,000 individuals across ethnicity^1–2^ and is caused by a loss of hypocretin/orexin in the hypothalamus, resulting in low/absent hypocretin (orexin) in the cerebrospinal fluid (CSF)^3^. Although the mechanism leading to hypocretin/orexin loss is unclear, it is likely autoimmune and involves genetic background and environmental factors, notably influenza.

Genome-wide association studies (GWASs) support autoimmunity. Loci include human leukocyte antigen (HLA)^4, 5^, T cell receptor (TCR) loci (both TRA and TRB)^6^, CTSH, IFNAR1, ZNF365, TNFSF4, CD207, NAB1, IKZF4-ERBB3, CTSC, DENND1B, SIRPG and PRF1^7^. Approximately 97-98% carry HLA-DQA1*01:02∼DQB1*06:02, a common haplotype encoding DQα*01:02 and DQβ*06:02, proteins that form the HLA-DQ0602 heterodimer, present in 12%–38% of controls^7^.

Interestingly, DQ0602 homozygous have a ∼2x increased risk, whereas DQ0602 heterozygous carrying DQα*01 or DQβ*05/06 (other DQ1) in *trans* of DQ0602 are at half risk. *Trans*-DQ1 heterodimers likely reduce DQ0602 amounts, a phenomenon we coined “allele” competition. This indicates that the amount of DQ0602 increases risk of occurrence, suggesting that an interaction of DQ0602 with specific antigen/TCRs is a bottleneck event in triggering autoimmunity. Similar doubling or more increases risk are reported in other diseases associated with single HLA molecules, such as celiac disease and DQA1*05/DQB1*02^8^, anti Lgi1 encephalitis and DRB1*07:01 ^9^ or spondyloarthropathies and B27^10^.

An unexplained finding in HLA analysis of susceptibility is that increased risk is also found in DQB1*06:02/DQB1*03:01 heterozygotes^7^. DQB1*03:01 was also found to affect disease onset in a Chinese study^7^. As DQB1*03:01 cannot heterodimerize with DQA1*01:02 for structural reasons^11^, the association is independent. Minor effects in other HLA loci are also present, including effects in DP and A^12^.

The major receptors of HLA peptide presentation are TCRs on T cells. Favoring a role of antigen presentation, narcolepsy is associated with polymorphisms in TCRA and TCRB. Functional analysis shows these affect TCR usage, decreasing TRAJ24, increasing TRAJ28, and reducing TRBV4-2 ^12^. The TRA SNP is also linked with a non-synonymous substitution in TRAJ24 (J24V->F) in the CDR3a region (position 116) that affects peptide recognition, although as mentioned this same variant (J24F) is expressed at lower level, thus this could also be involved.

Much progress has been made in understanding environmental events triggering narcolepsy^7; 13^. Pandemrix®, an ASO3 adjuvanted flu vaccine used in the 2009 pandemic H1N1 (pH1N1) epidemic in Europe, increased incidence in Northern Europe and other countries^14–16^. Incidence of T1N in China also increased 4 months after the H1N1 pandemic^17–19^, with weaker effects in US^20^, Taiwan^21^ and Germany^22^, countries where vaccination occurred at low rates. This suggests that flu epitopes favor an autoimmune reaction that escalates to hypocretin cell loss. Unfortunately, although increased T-cells reactive to pH1N1 and HCRT epitopes have been reported, differences in cell phenotypes and TCR usage that could link TCR use to genetic effects have not been found^23^, suggesting that an epitope remains to be discovered. Similarly, CD8+ T cells recognizing intracellular components of HCRT cells^24^ have been found, but no clear TCR usage preference observed.

Occurrence of narcolepsy shows a slight male preponderance. Onset of T1N is in childhood or early adulthood, although rare cases with onset at 2 or after 70 exist^7^. Interestingly, onset is younger in China and was reduced during the H1N12009 pandemic^17^. In this study we conducted a GWAS analysis of onset age, revealing a strong GWAS-significant effect of DQB1*03:01 across all ethnic groups.

## Results

### Age of onset across ethnic groups

Supplementary Figure 1A shows onset age distribution by ethnicity (defined by principal component analysis). Mean onset in East Asians (*n*= 2205) was 15.22 ± 10.98 years old (yo) while in Europeans (*n* = 1810), it was 21.12 ± 12.20 yo (*p*<0.01). In African Americans and European Africans, onset was 18.18±11.53 yo (*n*=178).

Most cases of East Asian origin were from China (n = 1748), Japan (n = 319) and Korea (n = 123). Using Analysis of Variance followed by post-hoc multiple (Dunnett’s T3), we found that mean onset age in Chinese (13.33 ± 10.28 yo) was lower than in Japanese (22.58 ± 10.78 yo, p < 0.01) and Koreans (22.83 ± 10.09 yo, p < 0.01), with no differences between Japanese and Koreans (p = 0.994). Cases of European origin were from the United States (21.53 ± 11.99 yo, n = 759) and Italy (21.47 ± 13.93 yo, n = 315) and did not differ in onset (p = 0.946).

### GWAS association of age of onset indicate a primary effect of DQB1*03:01

GWAS showed a signal within HLA-DQ in Caucasians (n = 1,797, rs41270875, P = 5.7×10^-10^) and East Asians (n = 2,201, rs17205738, P = 1.810^-9^) (suppl Figure 2A and C). Meta-analysis (n = 4,300) confirmed an association surrounding HLA DQB1 (rs34297496, P = 1.8×10^-15^) (Figure 1A and B, see Supplementary Figure 3 for ethnic specific plots).

**Figure 1.**
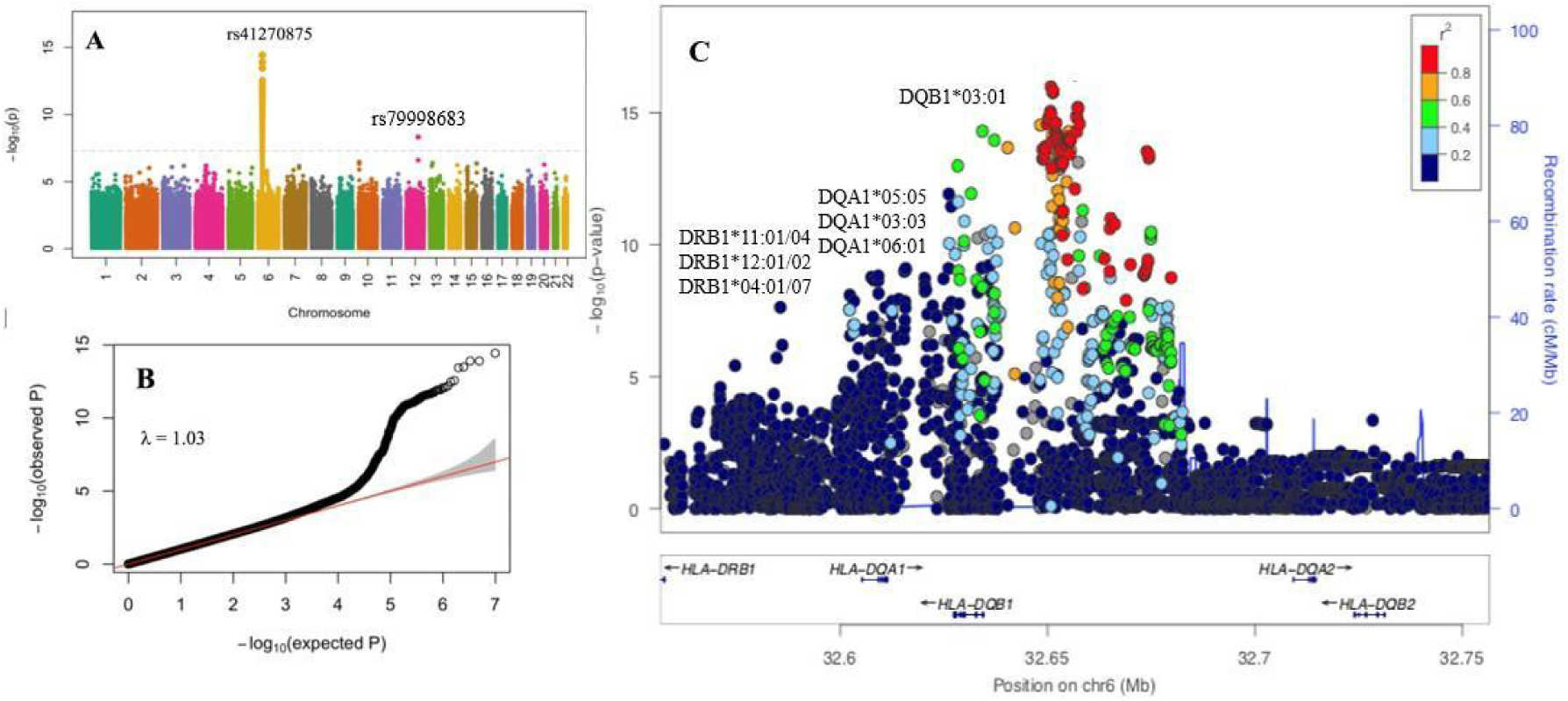
Genome wide association of narcolepsy age of onset. Manhattan plot of age of onset across ethnicity (A) with (B) QQ-plot and (C) zoom plot focused on the HLA DR, DQ region. Note largest effect surrounding DQB1, corresponding to shared DQB1*03:01, and highest p value for DQB1*03:01. A first drop in p value is observed telomeric in the region of DQA1, corresponding to DQA1*05:05, DQA1*03:03 and DQA1*06:01. A further drop is observed in the DRB1region, corresponding to the DRB1 region, where further haplotype diversity is observed. See supplementary Figure 2 for ethnic specific plots and more details on the DQB1 region of identity. Although rs79998683, another signal in chromosome 12 is significant, it is in a gene poor region and maps within long coding RNA ENSG00000257156.1

HLA alleles were next imputed^26, 27^. HLA-DQB1*03:01 was associated with onset across all ethnicities (P = 1.7×10-^18^, beta = −3.47 years (95%CI: −4.24, −2.70)) without heterogeneity. P values for associations in Caucasian, East Asians and others were 4.6×10^-10^ (beta = −3.90 (95%CI: −5.11, −2.68)), 7.8×10^-11^ (beta = −3.43 (95%CI: −4.45, −2.40)) and 0.7454 (beta = 0.62(95%CI: −3.10, 4.33)), respectively.

The associated GWAS zoom plot (Figure 1C) shows highest significance within DQB1, with a drop in DQA1 and DRB1, corresponding to known haplotypes containing DQB1*03:01 (see below). Supplementary Figure 3 shows zoom plots in Caucasians and Asians while Supplementary Figure 4 shows region of identity of the signal, which surround DQB1 exons.

Other HLA alleles with nominally significant associations are shown in Supplementary Table 1. Importantly, no effect was seen with DQB1*03:02 or DQB1*03:03, two frequent DQB1*03 alleles with modest divergence with DQB1*03:01 (6 and 5 amino acids respectively, see Supplementary Figure 6). Most notably, DQB1*03:02, a common allele in the context of DQA1*03:03∼DQB1*03:02 (typically DRB1*04 haplotypes) was not associated with age of onset.

The GWAS signal was next controlled for DQB1*03:01, showing disappearance of all signals (Supplementary Figure 5); inspection of remaining effects in each ethnic group (Supplementary Table 2), did not show any effect.

Association with DQB1*03:04 (DQB1*03:01-like except for 57 D>A, 3 cases) or DQB1*03:05 (DQB1*03:02-like, could not be imputed) could not be ascertained due to small numbers. Similarly, effect of DQB1*03:19 (13 cases) an African-specific DQB1* 03:01-like subtype was not significant (Supplementary Table 2; Supplementary Figure 6 for sequences).

### Effects of various DQB1*03:01 haplotypes on age of onset

DQB1*03:01 is found within haplotypes with distinct DQA1, DQA1*05:05, DQA1*03:03 (transethnic) and DQA1*06:01 (Asians). Further, DQB1*03:19 is present at moderate frequency in Africans in similar DR∼DQ haplotypes. Table 1 shows effects of DQA1∼DQB1 haplotypes on age of onset, while Supplementary Table 3 shows the same in extended DRB1∼DQA1∼DQB1 haplotypes. Remarkably similar effects are found for all DQB1*03:01-bearing haplotypes except perhaps in African Americans. In this ethnic group, the result is confounded by small sample size and the fact DQB1*03:01 is supplanted by DQB1*03:19, differing from DQB1*03:01 at position 185 (T>I)^26^. Insufficient power makes it difficult to establish whether DQB1*03:19 and/or DQB1*03:01 also reduces age of onset in this population.

**Table 1:** Effects of various DQA1-DQB1*03:01 haplotype on the onset age of narcolepsy in various ethnic groups.

| DQA1-DQB1 | *n* (with haplotype/without DQB1*03:01) | Mean age at onset (year, with haplotype/without DQB1*03:01) | BETA (95%CI) | *P* * |
| --- | --- | --- | --- | --- |
| All cases |  |  |  |  |
| 05:05-03:01 | 685/3125 | 15.99/18.71 | -3.38(-4.35, -2.41) | **1.10E-11** |
| 03:03-03:01 | 212/3125 | 17.49/18.71 | -3.55(-5.21, -1.88) | **3.17E-05** |
| 06:01-03:01 | 204/3125 | 11.76/18.71 | -4.37(-6.08, -2.67) | **5.54E-07** |
| Different Ethnicity |  |  |  |  |
| Caucasians |  |  |  |  |
| 05:05-03:01 | 362/1272 | 18.36/22.27 | -3.9(-5.35, -2.46) | **1.22E-07** |
| 03:03-03:01 | 170/1272 | 18.53/22.27 | -3.74(-5.72, -1.76) | **0.00022** |
| 06:01-03:01 | 0/1272 | - | - | - |
| East Asians |  |  |  |  |
| 05:05-03:01 | 288/1612 | 12.72/16.14 | -3.43(-4.82, -2.03) | **1.71E-06** |
| 03:03-03:01 | 40/1612 | 13.44/16.14 | -2.71(-6.26, 0.84) | 0.135 |
| 06:01-03:01 | 201/1612 | 11.75/16.14 | -4.39(-6.02, -2.76) | **1.44E-07** |
| Africans |  |  |  |  |
| 05:05-03:01 | 13/151 | 21.85/17.80 | 4.05(-2.33, 10.43) | 0.216 |
| 03:03-03:01 | 0/151 | - | - | - |
| 06:01-03:01 | 0/151 | - | - | - |
| 05:05-03:19 | 6/151 | 13.17/17.80 | -4.63(-13.58, 4.31) | 0.312 |
| Admixed |  |  |  |  |
| 05:05-03:01 | 21/78 | 14.81/15.90 | -1.09(-6.08, 3.89) | 0.668 |
| 03:03-03:01 | 2/78 | 10.50/15.90 | -5.40(-20.70, 9.89) | 0.491 |
| 06:01-03:01 | 3/78 | 12.67/15.90 | -3.24(-15.71, 9.23) | 0.612 |
| Different Nationality of East Asians |  |  |  |  |
| Chinese |  |  |  |  |
| 05:05-03:01 | 259/1231 | 11.96/14.07 | -2.11(-3.52, -0.70) | **0.003** |
| 03:03-03:01 | 30/1231 | 10.47/14.07 | -3.60(-7.46, 0.26) | 0.068 |
| 06:01-03:01 | 184/1231 | 11.12/14.07 | -2.95(-4.57, -1.32) | **3.9E-4** |
| Japanese |  |  |  |  |
| 05:05-03:01 | 17/276 | 20.29/22.83 | -2.54(-7.93, 2.86) | 0.358 |
| 03:03-03:01 | 9/276 | 21.83/22.83 | -1.00(-8.18, 6.19) | 0.786 |
| 06:01-03:01 | 4/276 | 16.50/22.83 | -6.33(-17.05, 4.39) | 0.248 |
| Korean |  |  |  |  |
| 05:05-03:01 | 11/94 | 19.36/23.53 | -4.17(-10.57, 2.24) | 0.205 |
| 03:03-03:01 | 1/94 | 27.00/23.53 | 3.47(-17.52, 24.46) | 0.747 |
| 06:01-03:01 | 10/94 | 20.60/23.53 | -2.93(-9.75, 3.89) | 0.401 |

\*P values calculated using generalized linear models in R. For “all cases”, p value was adjusted by ethnic group.

### Effects of DQB1*06:02 dosage on age of onset

Risk of narcolepsy is modulated by DQ0602 dosage through “allelic competition”. In this study, we found no effect of homozygosity or trans-competing alleles DQA1*01:03∼DQB1*06:01, DQA1*01:03∼DQB1*06:03 or DQA1*01:01∼DQB1*05:01 on age of onset even after controlling for DQB1*03:01 (Supplementary Tables 2 and 4).

### Effects of non-HLA-SNPs associated with narcolepsy occurrence on age of onset

Our recent narcolepsy GWAS identified not only DQ, but other loci^12^. Although no other peaks were found in our age of onset GWAS besides DQB1*03:01, we explored if these other loci could have effects on onset. As shown in Supplementary Table 5, little effect was found, although TRA rs1154155G, associated with narcolepsy, was nominally associated with an earlier age of onset.

### Binding motifs of DQB1*03:01 associated heterodimers

DQB1*03:01 comes in several heterodimeric DQA1 combinations, DQA1*03:03/DQB1*03:01 (Caucasians), DQA1*05:05/DQB1*03:01 (all ethnic groups) and DQA1*06:01/DQB1*03:01 (Asians). These alleles are divergent (Supplementary Figure 6 for sequences) but have resembling registers except for pocket 1 in DQA1*03:03/DQB1*03:01 where an acidic residue is preferred; DQA1*05:05/DQB1*03:01 and DQA1*06:01/DQB1*03:01 preferred Valine, Isoleucine or Proline (Supplementary Figure 7).

As controls, we also plot in Supplementary Figure 8 binding motifs of DQB1*03:02 and DQB1*03:03 alleles, alleles not associated with onset. These are found in DQA1*03:01/ DQB1*03:02, DQA1*02:01/DQB1*03:03 (DRB1*07:01-associated haplotype), and DQA1*03:02/DQB1*03:03 (DRB1*09:01-associated haplotype) heterodimers. As can be seen, resembling motifs are also seen, except for a Glycine at P4, which seems important in DQB1*03:01 associated alleles.

### HCRT epitopes binding DQB1*03:01 and DQB1*06:02

Although less probable, these results could suggest that a peptide binds DQB1*03:01 together with any DQA1 carrying a non-distinguishing residue in P1 and a G in P4 is involved. We therefore explored if HCRT sequences could bind DQA1*03:03/DQB1*03:01, DQA1*05:05/DQB1*03:01 and DQA1*06:01/DQB1*03:01. HCRT is a candidate autoantigen as increased frequency of T cells recognizing the N-terminal amidated end of hypocretin-1 has been reported^23; 27^ notably close to onset. Further, increased T cell reactivity to HCRT has been found by most^28–30^ but not all^31; 32^ investigators. HCRT has established DQ0602 binding motifs^28^.

Interestingly, binding for DQ0602^27^ and all three DQB1*03:01 heterodimers ranked high for RRCSAPAAASVAPGG and predicts the same <u>PAAASVAPG</u> core (Supplementary Table 6 and Luo et al^27^), although rank is lower for DQA1*03:03/DQB1*03:01 (rank: 0.19 vs 0.04 and 0.05) because Proline in position 1 is less favorable (Supplementary Table 4). This led us to hypothesize that binding of SA<u>PAAASVAPG</u>G to both DQ0602 and DQB1*03:01 could result in a younger age of onset through a competitive effect. A similar mechanism “epitope stealing” has been proposed to explain the DQ0602 protective effect in type 1 diabetes^33^, with InsB6-23 binding both DQ0602 and DQ8. In our case however, the common binder would need to synergize rather than compete, perhaps involving regulatory T cells.

To test this, we stained samples of DQ0602-positive controls and patients with this epitope using tetramers but found few positive cells even after a 10-day culture^23^. (Supplementary Figure 9). Considering this result, reactivity to the same peptide presented by DQB1*03:01∼DQA1 was not tested, as the hypothesis is unlikely.

### CDR amino-acid TCR differences in DQB1*03:01 vs other

TCRs interact with HLA independent of peptide presentation, such that this results in usage preferences in the presence of specific HLA subtypes, the result of positive selection in the thymus. In other words, HLA polymorphisms are trans-QTL of TCR usage^34–36^, and this occurs independently of DQA1 (and vice versa). Considering the strong association of narcolepsy occurrence with TCRA and B polymorphisms modulating the TCR repertoire, notably TRAJ24, TRAJ28 and TRBV4-2, an effect of DQB1*03:01 on the TCR repertoire could also modulate disease onset.

As TCR binds HLA at an angle^34; 35^, interactions with DQB1 occur at the level of TCR CDR1/2a on the mouth of the HLA binding cleft^34–36^ when using the preferred peptide docking forward alignment (P1–P9 from N- to C-terminus) of most HLA-TCR crystal structures. Interaction of DQB1 with CDR1/2b is also possible in the alternate alignment. More recently, strong and complex effects have been reported within the CDR3 TCR hypervariable region^34^, perhaps through direct contacts and indirectly through peptide binding, or through CDR3 interactions with the CDR1 and 2 loops mentioned above that themselves interact with HLA.

To address this, we used three datasets, one with Caucasian data and bulk RNA TCRA and TCRB sequences (DGN), and two single cell data (CHINA and CAUC) (see Methods and Supplementary Data). Because bulk DGN sequencing data does not contain CDR3 data, we mostly used single cell data (CHINA and CAUC) to compare TCR repertoire. The bulk DGN data was only used as additional confirmation for detection of CDR1a and CDR2a effects.

A strength of this approach is use of two techniques and two ethnic groups (CHINA and CAUC). Because DQB1*03:01 associated DQA1 alleles have different distributions in Asians versus Caucasians, differences showing no heterogeneity will be robust, and specific of DQB1*03:01, independent of DQA1 (or DRB1). CDR amino acid positions within TCR sequences were encoded as per the international ImMunoGeneTics information system (IMGT, https://www.imgt.org/IMGTindex/TR.php) for CDR1 and CDR2 as and as per Hishigaki et al^34^ for CDR3.

Results of single cell analysis in Caucasians and Chinese are reported in Figure 2, with star (*) indicating position/amino acid differences found to be similarly significant across ethnic groups (see also Supplementary Table 7).

**Figure 2.**
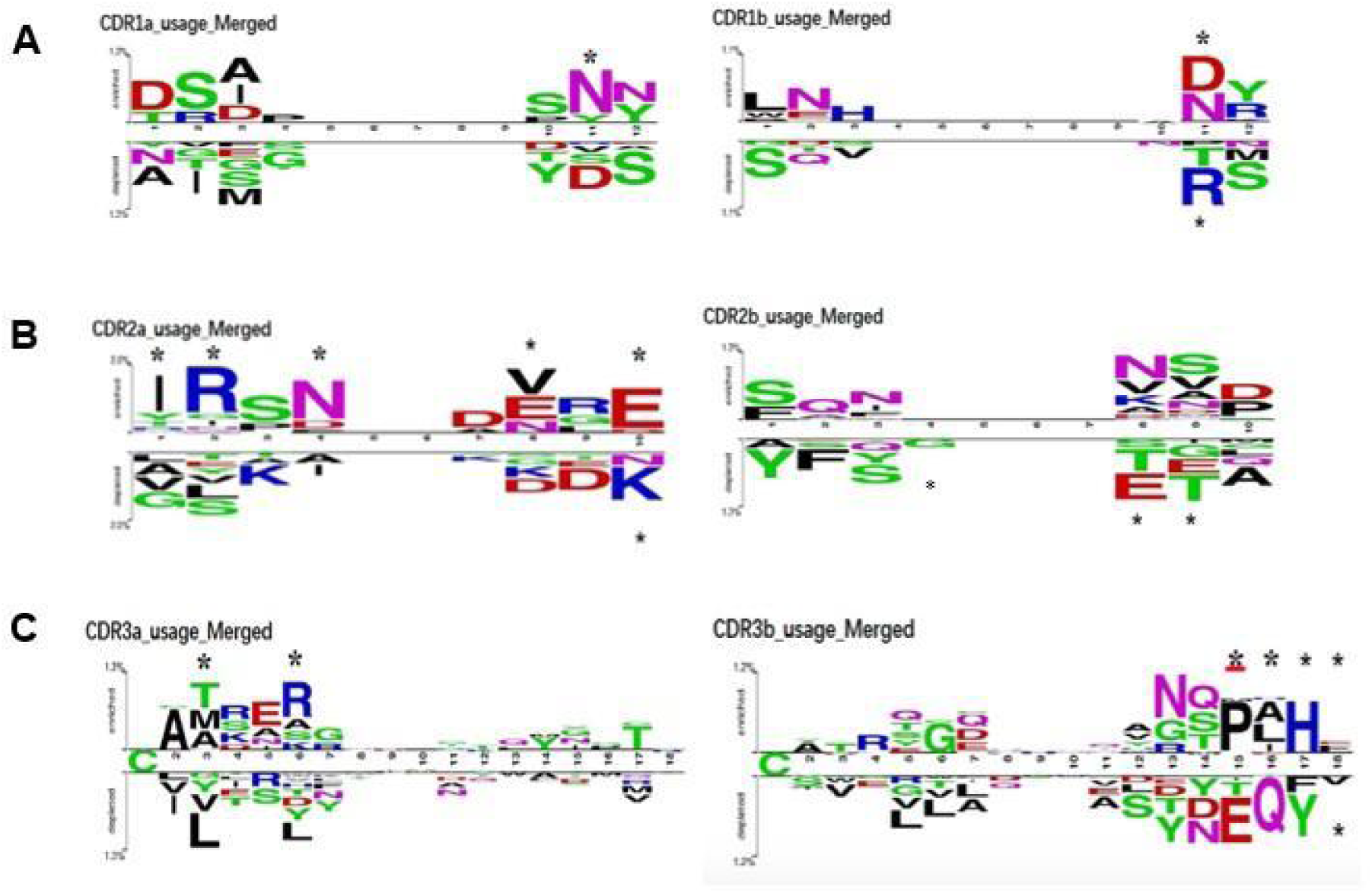
Aminoacid usage preference with TCRA and B CDR sequences between DQB1*03:01 positive and negative subjects. Data was derived from single cell sequencing in Chinese (CHINA) and Caucasian (CAUC) subjects (see methods for datasets). For simplification, usage preference (% usage of aminoacid in positive or negative deviation) is displayed with equal representation of Chinese and Caucasian data. A: CDR1 alpha (right) and beta (left); B: CDR2 alpha (right) and beta (left); C: CDR3 alpha (right) and beta (left). Position with significant findings are denoted with a star. For exact p values for each aminoacid, see supplementary tables 7-11.

As can be seen, the most significant differences across ethnicity (thus independent of DQA1) were a preference for R at position 2 of CDR2a, an absence of T at position 8 of CDR2b and presence of F and not V at position 18 of CDR3b. Most differences were in CDR2a, where a motif IR-N was found, reflecting increased usage of TRAV13-1, TRAV17 and TRAV23/DV6. It is notable that TRAJ24 CDR3a F/L at position 17 (genetically associated with narcolepsy) was not over or underrepresented. Similarly, amino acid characteristics of TRAJ28 are not present and TRBV4-2, which has a favorable N at position 11 of CDR1b, has no favorable residue within CDR2b. Overall, known genetically associated TRAJ and TRBV polymorphisms are not particularly likely to interact with DQB1*03:01.

## Discussion

In this study, we report on a novel, robust, transethnic association of DQB1*03:01 with earlier age of onset. This association is indistinguishable to DQB1 amino-acid substitution 45 G to E, a polymorphism shared by DQB1*03:01 and DQB1*03:19, an African American subtype.

The DQB1*03:01 effect is independent of sex or age of onset itself. Rather, onset is shifted across all ages (Supplementary Figure 1) and does not interact with puberty. This association is significant in Asians and Caucasians without heterogeneity, even so age of onset is lower in Chinese. Whether or not the younger age of onset in China reflects differences in genetics, ascertainment or exposure to triggering events is unknown.

Interestingly, in Africans, association is opposite, DQB1*03:01-positive having a non-significant higher age of onset (Table 1). Although this is likely due to the small number of cases (13 in 151 cases), it could also be due to the special clinical presentation reported in this group where more cases with hypocretin deficiency and without cataplexy have been found^25^. Further, in cases with hypocretin deficiency (low hypocretin) but without cataplexy, only half are estimated to ever develop cataplexy^38^.

Atypical presentation of African Americans may interact with DQB1*03:01 differently, making the effect harder to detect. Increased sample size, with confirmation of DQB1*03:01 versus DQB1*03:19 status in this ethnic group will be needed to distinguish these possibilities.

Strikingly, DQB1*06:02 homozygosity increases risk of narcolepsy^12^ but has no effect on onset (Supplementary Table 4) or severity^5^. This indicates a differential regulation of onset versus risk. Similarly, other SNPs associated with narcolepsy risk have minor effects on age of onset, although TRA rs1154155 G, associated with narcolepsy, decreases onset age by 1 year, with nominal significance (Supplementary Table 5). It is our hypothesis that the amount of DQ0602 heterodimers regulates probability of a chance encounter with a rare autoantigen that triggers the initial process, while DQ0301, together with other polymorphisms have downstream effects leading to the full-blown syndrome. These results suggest that effects regulating the probability of developing the disease and how rapidly/completely the disease manifests may be distinct.

A limitation may be ascertainment biases, as there are arguments suggesting that a portion of patients with hypocretin deficiency and no cataplexy, representing a less severe phenotype, may be less frequently diagnosed. Indeed, in families of patients with narcolepsy-cataplexy, 1% of first-degree relative show narcolepsy-cataplexy, but a similar or higher percentage of patients (1-2%) also report narcolepsy-like symptoms but no cataplexy, sometimes in the presence of hypocretin deficiency, with suggestion that the loss of hypocretin neurons may not be as complete. It may therefore be that cases with hypocretin deficiency without cataplexy are not uncommon in the general population but mild enough not to come to the attention of sleep clinics. This is supported by one study where sleep studies were conducted in ∼1000 subjects of the general population and where one-two narcolepsy without cataplexy case and one case with cataplexy were identified. In this context, presence of DQB1*03:01 or other polymorphisms could favor a more rapid and complete destruction of hypocretin cells. Only once a blood test will be available for hypocretin deficiency or for autoimmunity (for example an autoantibody) will the true spectrum of the disease in the general population be ascertainable.

Considering this, we next explored mechanisms. Indeed, across three DQA1∼DQB1*03:01 haplotypes, the region of identity surrounds DQB1*03:01. One possibility could be that a regulatory polymorphism (Supplementary Figure 4) is involved, for example modulating DQ0602 presentation in *trans* of DQB1*03:01 (almost all subjects are DQB1*06:02). Such polymorphisms have been reported by Raj et al^38^ and Kang et al^39^, although regulations are *cis* located, thus would not affect DQB1*06:02. Weiner-Lachmi et al^40^ explored if DQB1*03:01 in *trans* of DQ0602 modulate DQB1*06:02 expression and heterodimer abundance on the surface of antigen-presenting cells and found no effect. This contrasted with DQ0602 homozygosity that doubled abundance. This result, in line with the lack of effect of DQ0602 homozygosity on onset (Supplementary Table 4), suggest that *trans*-modulating regulatory elements on DQB1*06:02 expression are unlikely to be involved.

Another possibility could involve peptide binding of an important autoimmune epitope across DQB1*03:01 and DQ0602 heterodimers. A similar mechanism has been postulated to mediate resistance effects of DQ0602 in diabetes through “epitope stealing”^33^, although modulation of the TCR repertoire including regulatory T cell^41^ has also been proposed. In narcolepsy, DQB1*03:01 *reduces* onset independently of DQA1, making epitope stealing unlikely. Nonetheless, we used peptide binding prediction for all three DQB1*03:01 heterodimers and found motifs to be similar except for P1 (Supplementary Figure 7). At this position, DQA1*03:03/DQB1*03:01 prefers an acidic residue whereas DQA1*05:05/DQB1*03:01 and DQA1*06:01/DQB1*03:01 prefer Valine, Isoleucine or Proline (Supplementary Figure 7).

In comparison, binding motifs for DQA1*03:01/DQB1*03:02, DQA1*0201/DQB1*03:03 and DQA1*0302/DQB1*0303, subtypes with no effect on narcolepsy onset, have variable motifs, although that of HLA-DQA1*0302/DQB1*0303 resemble DQA1*03:03/DQB1*03:01 (Supplementary Figure 8). Overall, these results show that motifs across DQB1*03:01 heterodimers, although similar, are not much more different between them than with DQB1*03:02 or DQB1*03:03 that are not associated with narcolepsy. We thus believe this hypothesis to be unlikely.

Pursuing on this, we nonetheless screened HCRT for DQ0602 binders that can bind all three DQB1*03:01 heterodimers. In doing so, we identified RRCSA<u>PAAASVAPG</u>G, predicted to bind DQ0301 and DQ0602 with PAAASVAPG (Supplementary Table 6). Nonetheless, rank was lower for DQA1*03:03/DQB1*03:01 (rank: 0.19 vs 0.04 and 0.05) because Proline in position 1 is less favorable to this heterodimer (Supplementary Figure 7 and Supplementary Table 6). Further, a similar core would be predicted with DQA1*03:03/DQB1*03:02, a common subtype that does not modulate onset. Finally, DQ0602-RRCSA<u>PAAASVAPG</u>G tetramer staining in PBMCs did not show abundant or disease-specific staining (Supplementary Figure 9).

Considering the above, a more likely explanation is a direct effect of DQB1*03:01 which favors selection of TCRs involved in the disease, perhaps downstream of the initiating event. As mentioned above, a striking feature of narcolepsy is its association with TCR polymorphisms that modulate usage of TRAJ24, TRAJ28 and TRBV4-2. Unfortunately, however, published work has not found effects of DQB1*03:01 on TCR segment usage or CDR3 diversity^34–36^, although focus was not on this HLA subtype. Similarly, here we found no effects of DQB1*03:01 on TRAJ24 and TRAJ28 usage, although a slight increase in TRBVB4-2 usage was observed, reflected by CDR1a/b interactions. This is also in agreement with the fact TCR polymorphisms associated genetically with narcolepsy do not have strong effects on age of onset (Supplementary Table 5). Clearly, in this disease, onset age and predisposition are regulated by different HLA subtypes and mechanisms.

Pursuing on this, we sought to identify DQB1*03:01-specific effects on TCR repertoire across ethnic groups, finding significant amino acid differences located across multiple regions of TCR CDRa and b loops.

Interestingly, highest preferential positions were a R at position 2 of CDR2a, an absence of G and T at position 4 and 8 of CDR2b respectively, and a preference of F vs V at position 18 of CDR3b. The CDR2a finding of preferential IR_N, one of the strongest findings, also replicated in a third Caucasian bulk sequencing dataset (DGN, see Supplementary Table 9). The stronger preferential (increased) amino acid usage found within the CDR1a (N11) and CDR2a loops (IR-N at P1,2,4), are in line with interactions of CDR1/2a and b with DQB1 around positions 70 to 80 as found in known structures^42–47^. In addition, strong effects in CDR3a and CDR3b usage were found, involving hydrophobic residues and histidine at P15-18. This is in line with Ishigaki et al^34^ who found that diversity within CDR3 TCR regions is modulated by HLA polymorphisms, notably amino acid 71 within DQB1. Although these effects are difficult to interpret considering state of knowledge of HLA-peptide-TCR involvement in narcolepsy, they represent complementary data for our understanding on how TCR-HLA interactions modulate narcolepsy risk.

Although these differences are notable, these do not easily explain how the only difference between DQB1*03:01 and all other DQB1 subtypes (including DQB1*03:02 and DQB1*03:03), amino acid 45G>E, could be involved. Indeed, the 45G>E substitution is located on the side of HLA-DQ, outside of the typical HLA-TCR interaction area^34–36^. It is notable for a change from a small glycine in all other DQB1 to a large, steric, and charged aspartic acid, which creates a polar acidic protrusion of ∼5 Å on the side of the usual TCR-HLA interacting area, on top of a secondary minor side ridge of the DQ molecular structure. Although speculative, atypical interactions with TCR involving R in CDR2a could be involved, although this would involve displacement of the CDR2a loop outside of its normal configuration, perhaps in the presence of a large CDR3b loop. Residues handing outside of the core binding of a peptide such as a glycosylated residues could also be involved. Crystal structures involving DQB1*03:01-TCR molecules are not available to explore how 45G>E affects HLA-TCR binding. Alternatively, and perhaps more likely, DQB1*03:01 effects may be modulated by multiple amino acid differences between this and other DQ subtypes and not a single amino acid.

Although these predictions are in line with structures of TCR-HLA interactions which involve CDR2α interactions with DQβ in the classical TCR binding configuration, the amino acids involved do not fit well. Indeed, in the classical model, supported by structural studies and trans QTL effects^35; 36^, CDR2α amino acids interact with DQB1 are CDR2 29, 31, 50 and 51 and those primarily interact with HLA-DQB1 amino acid 66-73, on the mouth of the HLA binding cleft. Problematically, however, these DQB1 66-73 amino acids are identical across all DQB1*03 subtype, making this unlikely to mediate this difference. Further, CDR2α I36 and R37 are not CDR2α amino acids known to support this interaction.

CDR1-CDR3 interactions with HLA-DQB1*03:01 could also be involved, although effects observed in other loops were far less significant than CDR2α 38R. The fact that increased usage of TRBV4-2 was found in DQB1*03:01 subjects could also be involved, an effect mediated by CDR1α. In our GWAS, a SNP associated with narcolepsy increases usage of TRBV4-2. Unfortunately, however, this TCRb polymorphism does not modulate age of onset, making this unlikely.

## Materials and Methods

### Study subjects

Subjects have been described in Ollila et al (2023)^12^. Cases (n=5339) were DQB1*06:02 and with clear-cut cataplexy or had narcolepsy with low CSF hypocretin-1. Age of onset of cataplexy and excessive sleepiness was found in 4450 cases, and the younger age of the two used as “age of onset”.

### Datasets used for TCR usage and CDR amino-acid analysis

Three datasets (DGN, CHINA, and CAUC) were used (see supplementary material), two (DGN and CHINA) for CDR1-2 and segment usage analysis, and two for CDR3 analysis (CHINA and CAUC).

### TCR amino acid and segment usage in the presence of DQB1*03:01

For bulk sequencing in Caucasians (DGN), we constructed a combined data matrix with each DQB1*03:01 genotype and TCR V, J usage. Python was used to build an ordinary least squares (OLS) model that predicted usage by DQB1*03:01 status. V and J segments for which usage was predicted by DQB1*03:01 with p < 0.05 after FDR correction were retained. We also report mean usage within DQB1*03:01-positive (n=306) and negative (n=589) individuals (Supplementary Table 8). In addition, known V segment sequences were used to build an aminoacidic matrix of position location within CDR1 and CDR2 loops. The symbol “.” was used for positions where no amino acid was present. Another OLS model was used to predict amino acid usage at each position stratified by DQB1*03:01 (Supplementary Table 9).

In the scRNA-seq datasets (CHINA), counts for each segment usage per cell was available. To verify adequacy, we correlated usage of all V and J segments of TCRA and TCRB across bulk DGN and single cell CHINA/CAUC datasets, finding consistent usage across methodologies. Next, we built 2×2 contingency tables for each segment (i.e. TRAV4 and non-TRAV4 counts in DQB1*0301-positive and negative) and used the chi2_contingency function from scipy.stats in Python to compare groups. FDR correction was performed and p-values < 0.05 used as significant. We also report odds ratio from each test (Supplementary Table 10). The same chi-squared model was built to test significance of interaction between DQB1*03:01 and each TCR amino acid, with “.” being used for positions with no amino acid (Supplementary Table 11).

### Genome wide association study and HLA imputation

Cases were genotyped using Affymetrix and analysis performed as in supplementary material and Ollila et al. 2023^12^. HLA imputation was performed using HIBAG^25^ (http://cran.r-project.org/web/packages/HIBAG/index.html). Imputed alleles with a posterior probability less than 0.4 were excluded as in Khor et al.^15^; only 17 cases were excluded.

### Analysis of HLA variants

To analyze effects of HLA on age of onset, generalized linear models were used, with control of ethnicity. Presence versus absence of each allele was compared, and, as almost all but a handful of subjects are DQB1*06:02, this corresponded to analyzing effects of the DQ haplotype located in *trans* of DQA1*01:02∼DQB1*06:02. Once association with DQB1*03:01 was established, additional associations were explored after conditioning or excluding DQB1*03:01-positive patients. Similar analyses were done in subjects with individual DQA1∼DQB1*03:01 or DRB1∼DQA1∼DQB1*03:01 haplotypes.

### Tetramer staining

DQ0602-SAPAAASVAPGG tetramers were built by peptide exchange and used to stain Peripheral Blood Mononuclear Cells (PBMCs) by FACS as described in Luo et al ^23^.

## Conclusions

In summary, we found a strong DQB1*03:01 effect on age of onset best explained by a DQB1 substitution of amino acid 45G to 45E. Explorations of mechanisms suggest peptide-binding independent effects on the TCR repertoire. Conserved effects across ethnicity allowed unique genetic mapping, which in time will provide a specific explanation.

## Data Availability

All data produced in the present study are available upon reasonable request to the authors

## Acknowledgments

We are indebted to all participants of the study, most notably narcoleptic patients. The study was partially funded by NIH AI144798.

---

Supplementary Figure 1: Age of onset distributions by ethnicity and DQB1*03:01 status

**Supplementary Fig. 1A.**
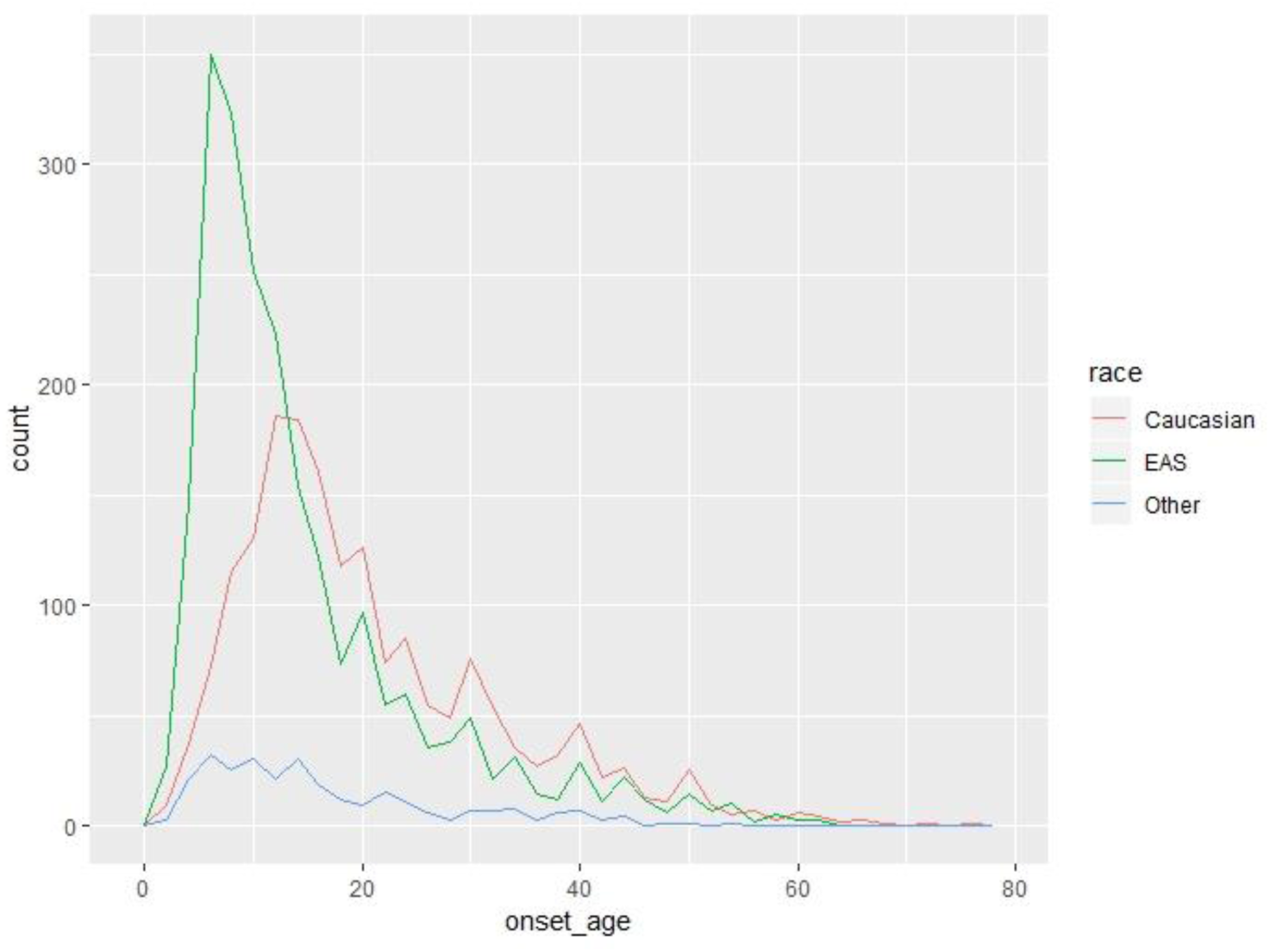
Distribution of onset age of narcolepsy across different ethnic groups (Causasian; EAS=East Asians; Other=mostly African Americans)

**Supplementary Fig. 2A.**
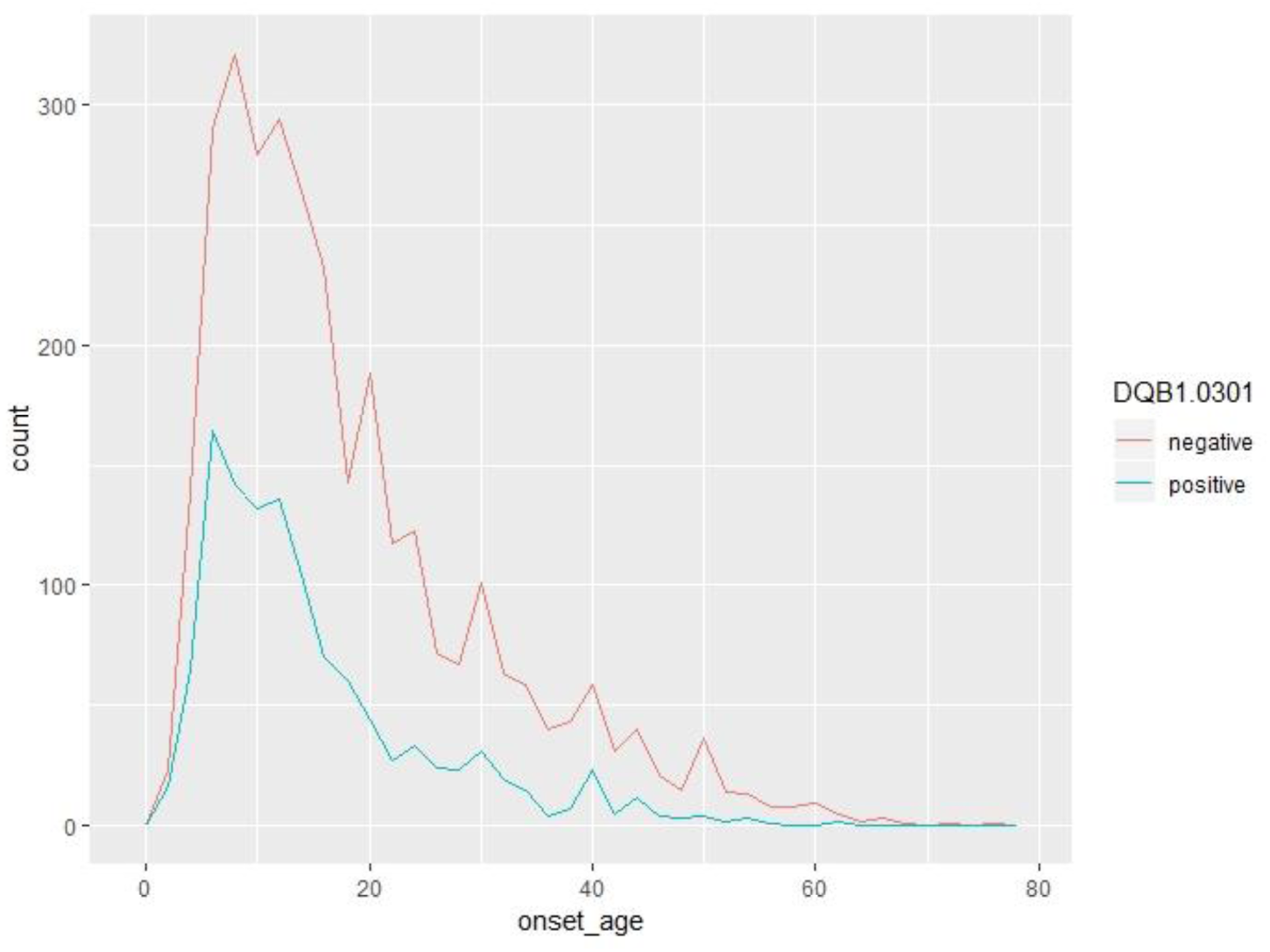
Distribution of onset age of narcolepsy in all HLA-DQB1*03:01 positive vs negative cases.

**Supplementary Figure 1C.**
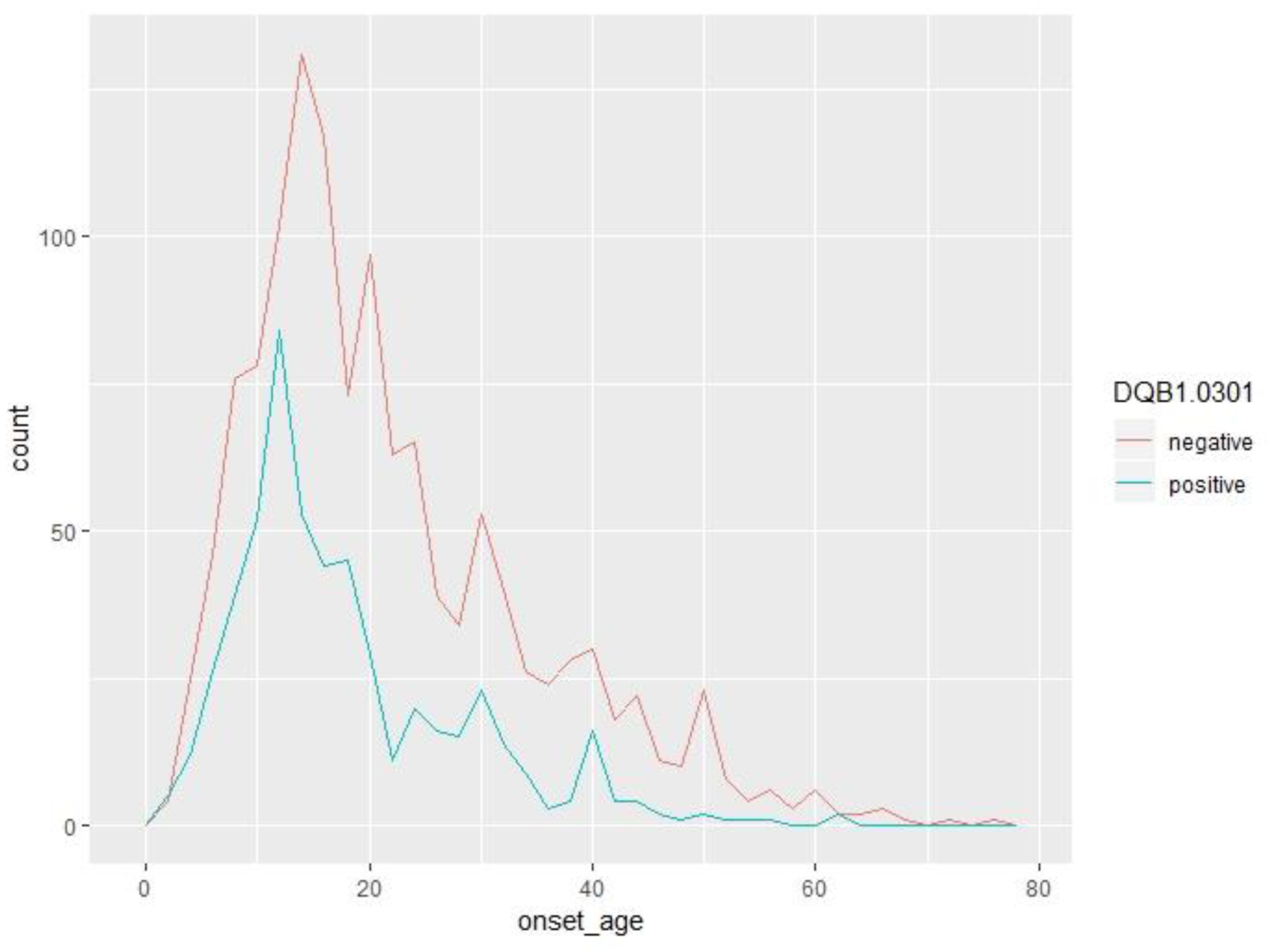
Distribution of onset age of narcolepsy with HLA-DQB1*03:01 positive or negative in Caucasian cases

**Supplementary Figure 1D.**
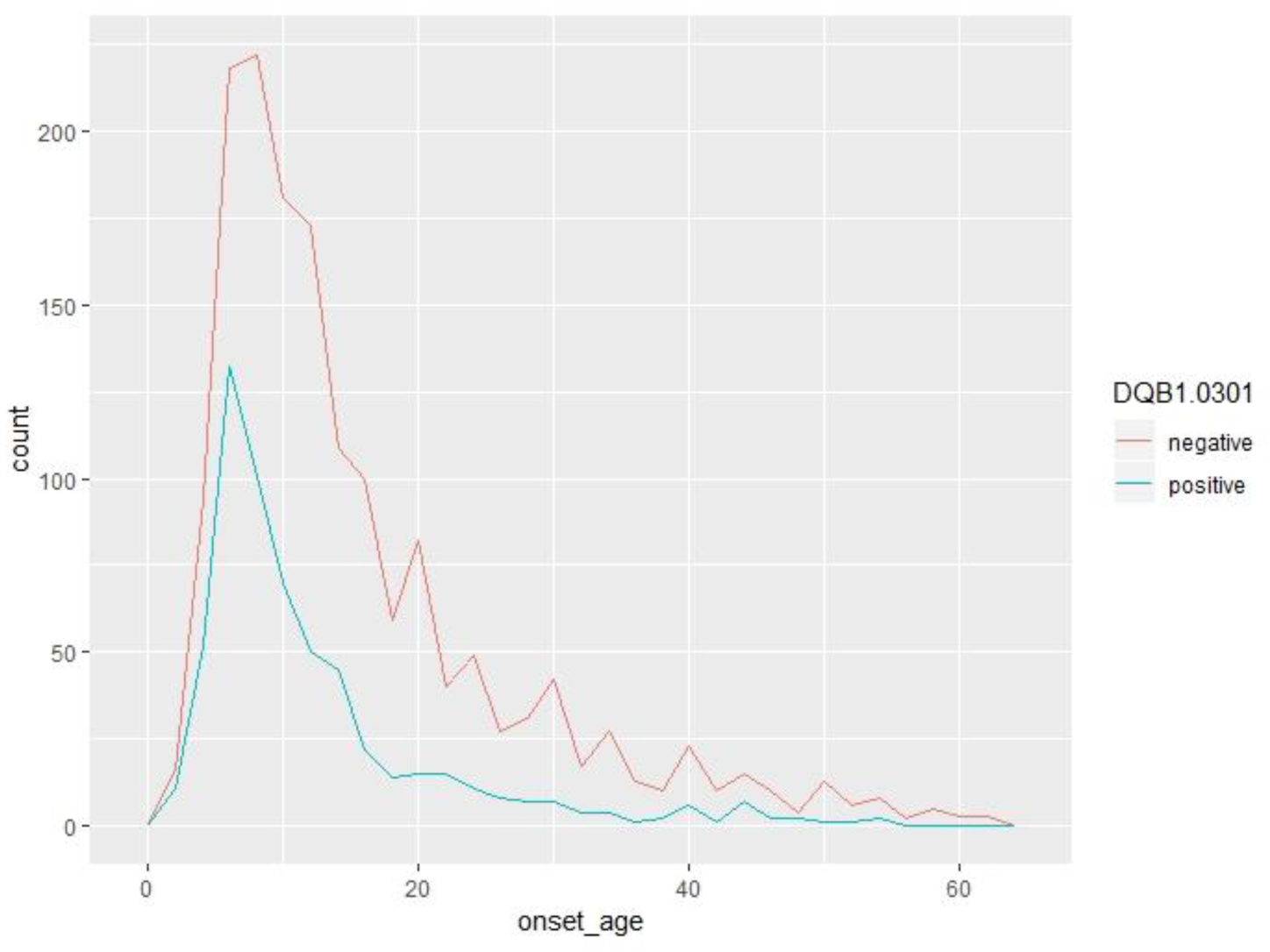
Distribution of onset age of narcolepsy with HLA-DQB1*03:01 positive or negative in Asian cases

**Supplementary Figure 1E.**
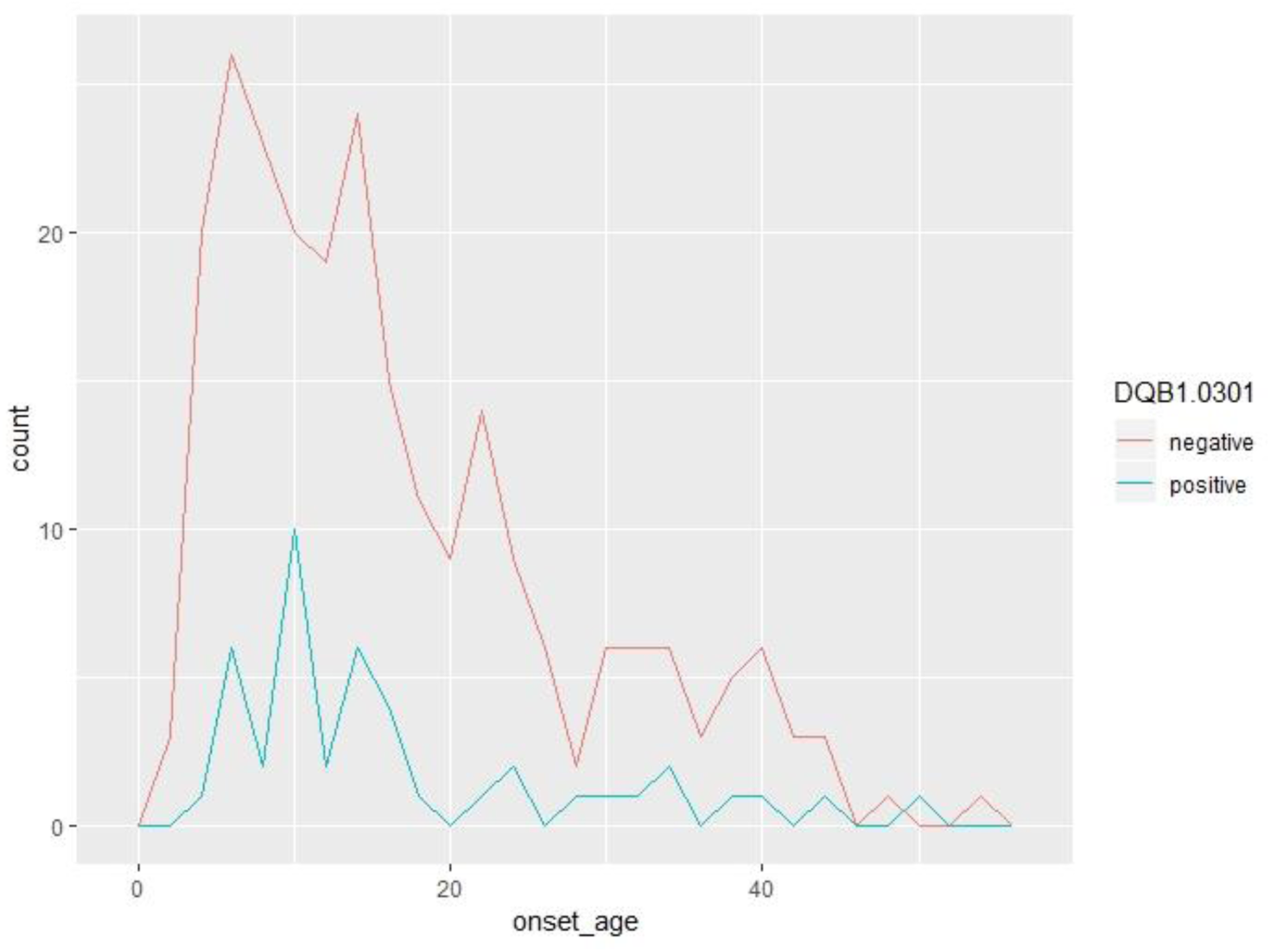
Distribution of onset age of narcolepsy with HLA-DQB1*03:01 positive or negative in other cases (AFR plus Admixed)

**Supplementary Figure 2:**
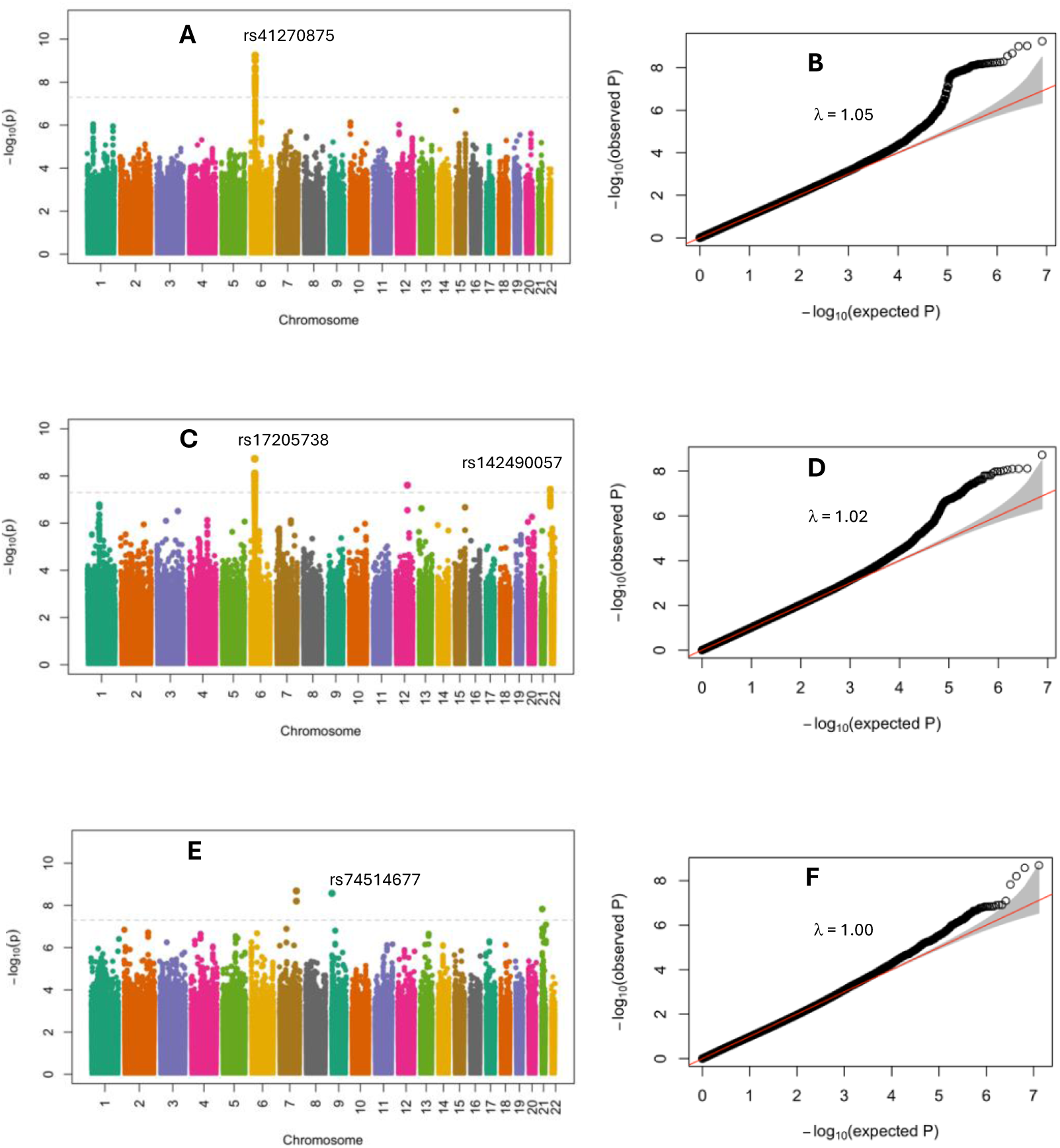
Age of onset GWAS by ethnic group. Manhattan Plot and QQ Plot of age at onset of narcolepsy. A, C, E represent the Manhattan Plots of Caucasian (n = 1797), East Asian (n = 2201), and AFR (n = 191), respectively. B, D, F represent the corresponding QQ Plots. rs142490057 is significant in East Asians and maps within the cat eye syndrome chromosome region (CECR) cluster that contains Adenosine Deaminase 2 (CECR2), a gene which when mutated is associated with neutropenia. rs74514677 is significant in Africans and locates close to INSL4 (insulin-like 4 protein or early placenta insulin-like peptide) a gene with restricted expression in human placenta. These other signals would need confirmation and were ethnic specific.

**Supplementary Figure 3:**
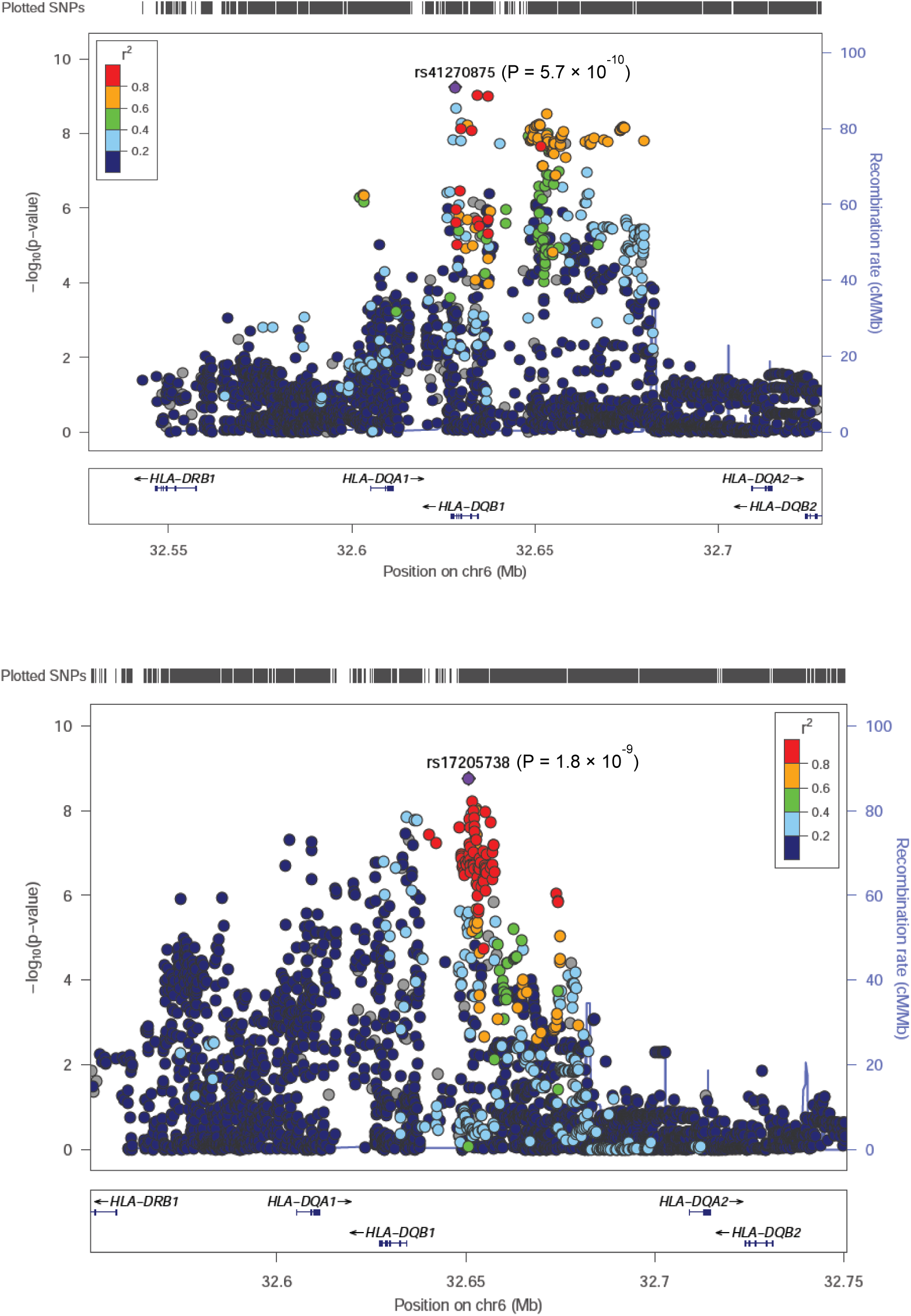
Zoom plots of HLA region in Caucasians (A) and East Asians (B).

**Supplementary Figure 4:**
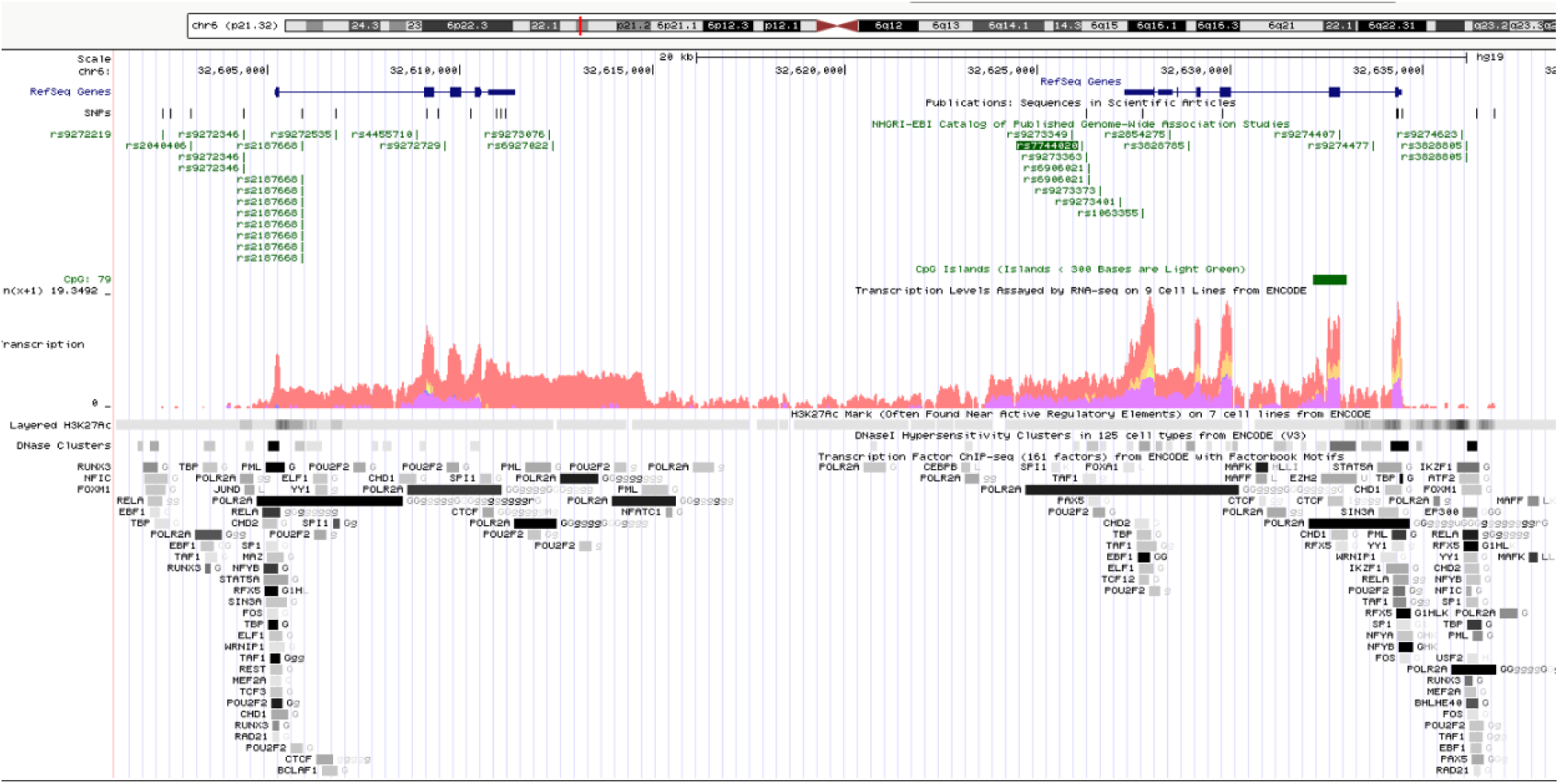
DQB1*03:01 Genomic segment associated with disease onset and associated regulatory elements. Note that elements in the region have been shown to regulate HLA expression, although none map in high LD with our lead SNPs, which track DQB1*03:01.

**Supplementary Figure 5:**
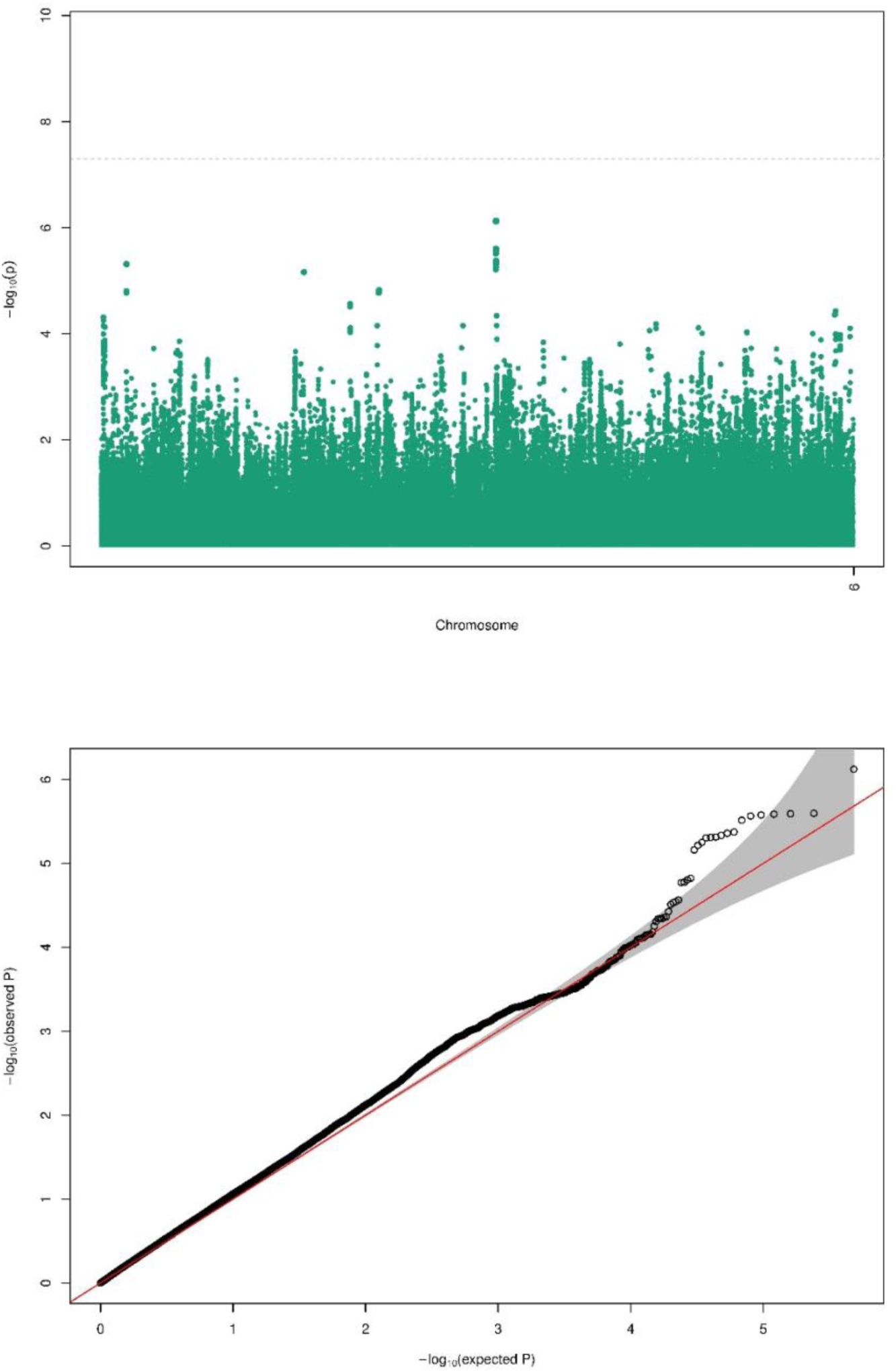
Disappearance of the chromosome 6 GWAS signal after controlling for DQB1*03:01

**Supplementary Figure 6:**
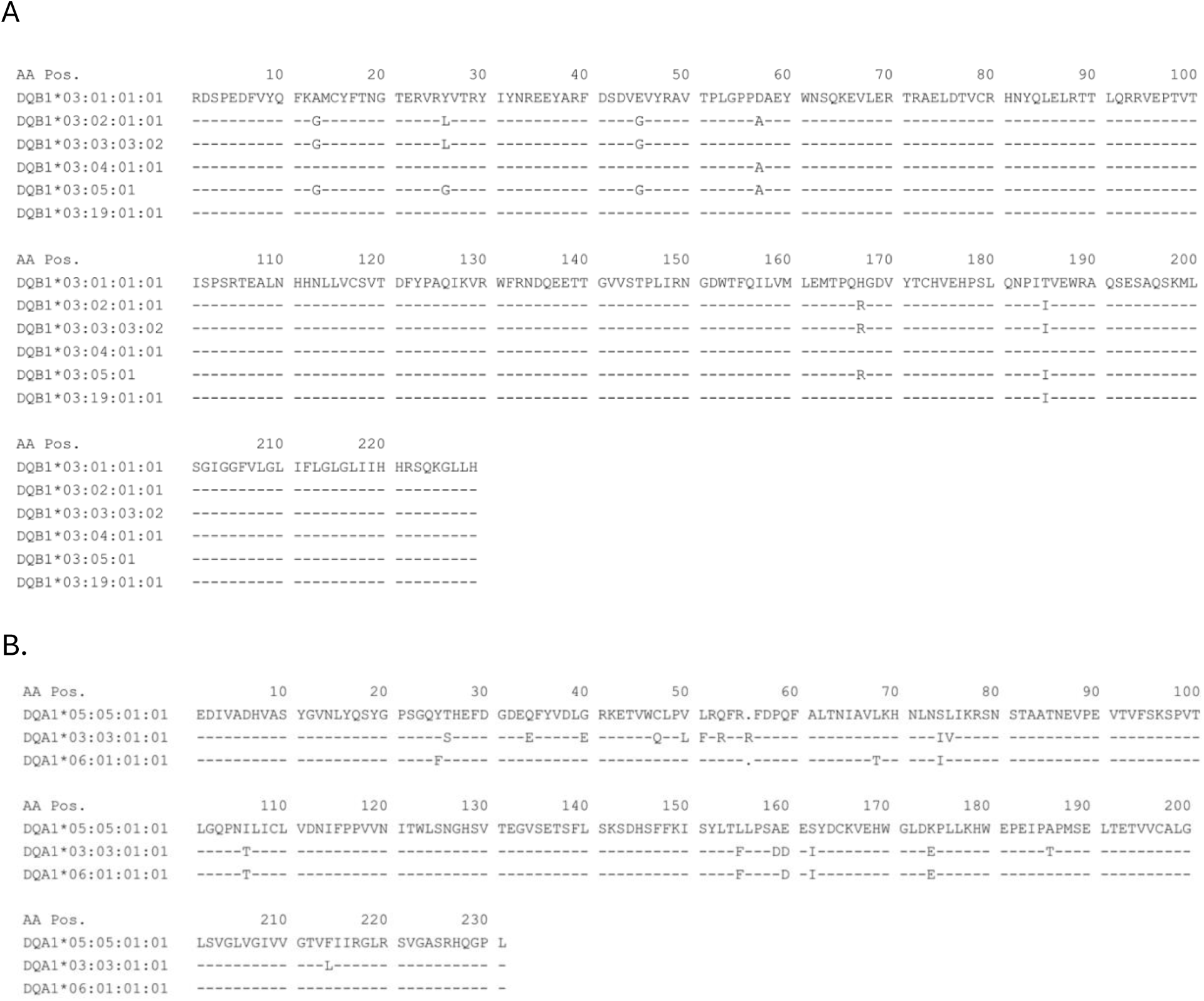
A. Sequence alignment of classic DQB1*03 alleles in comparison to DQB1*03:01. DQB1*03:02 and DQB1*03:03 are extremely common alleles, and these are clearly not associated with younger age of onset. DQB1*03:04 and DQB1*03:05 are very rare and could not be tested. DQB1*03:19 is reasonably common in Africans but is not found in any other ethnic groups. Because our African sample is small, it was not possible to test if this allele is associated with a younger age of onset. Of note, aminoacid E45 (instead of G45) is the most specific marker of DQB1*03:01, also shared by DQB1*03:19 but not present in any other DQB1 subtypes; it is highly linked with our signal r2∼1.0. B. Alignment of DQA1 alleles commonly associated with DQB1*03:01. Note the very large sequence diversity.

**Supplementary Figure 7:**
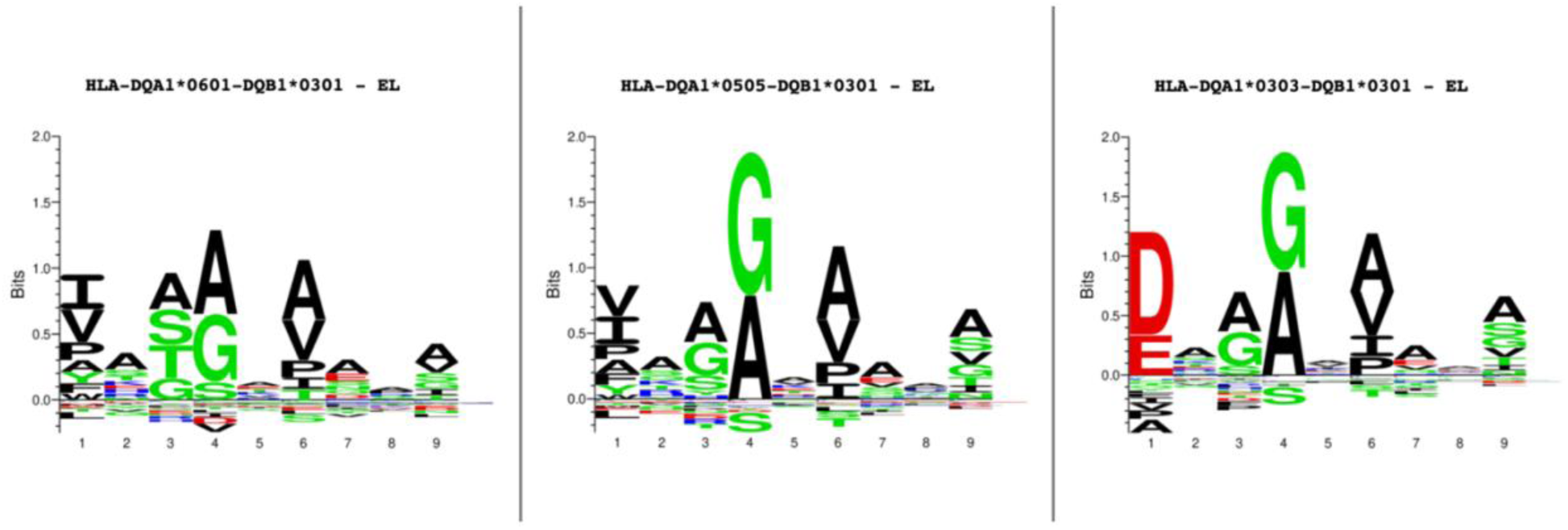
HLA binding motifs of 3 DQA1∼DQB1*03:01 heterodimers

**Supplementary Figure 8:**
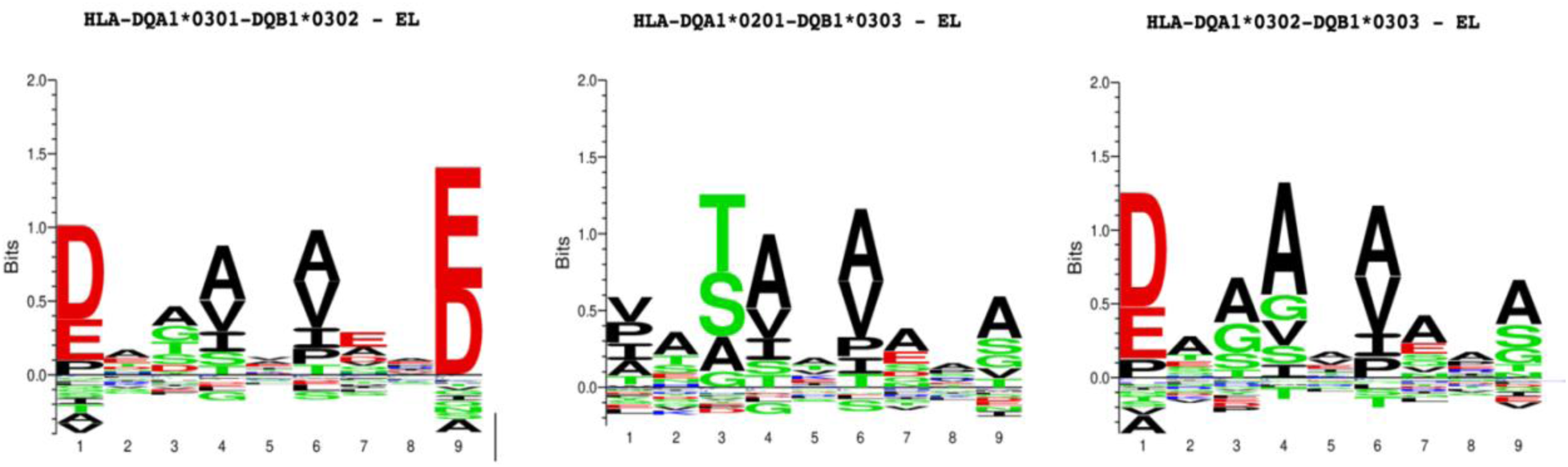
HLA binding motifs of 3 common DQA1∼DQB1*03 heterodimers (DQA1*02:01/DQB1*03:03 (DRB1*07:01 associated haplotype), DQA1*03:02/DQB1*03:03 (DRB1*09:01 associated haplotype) or DQA1*03:03/DQB1*03:03.

**Supplementary Figure 9:**
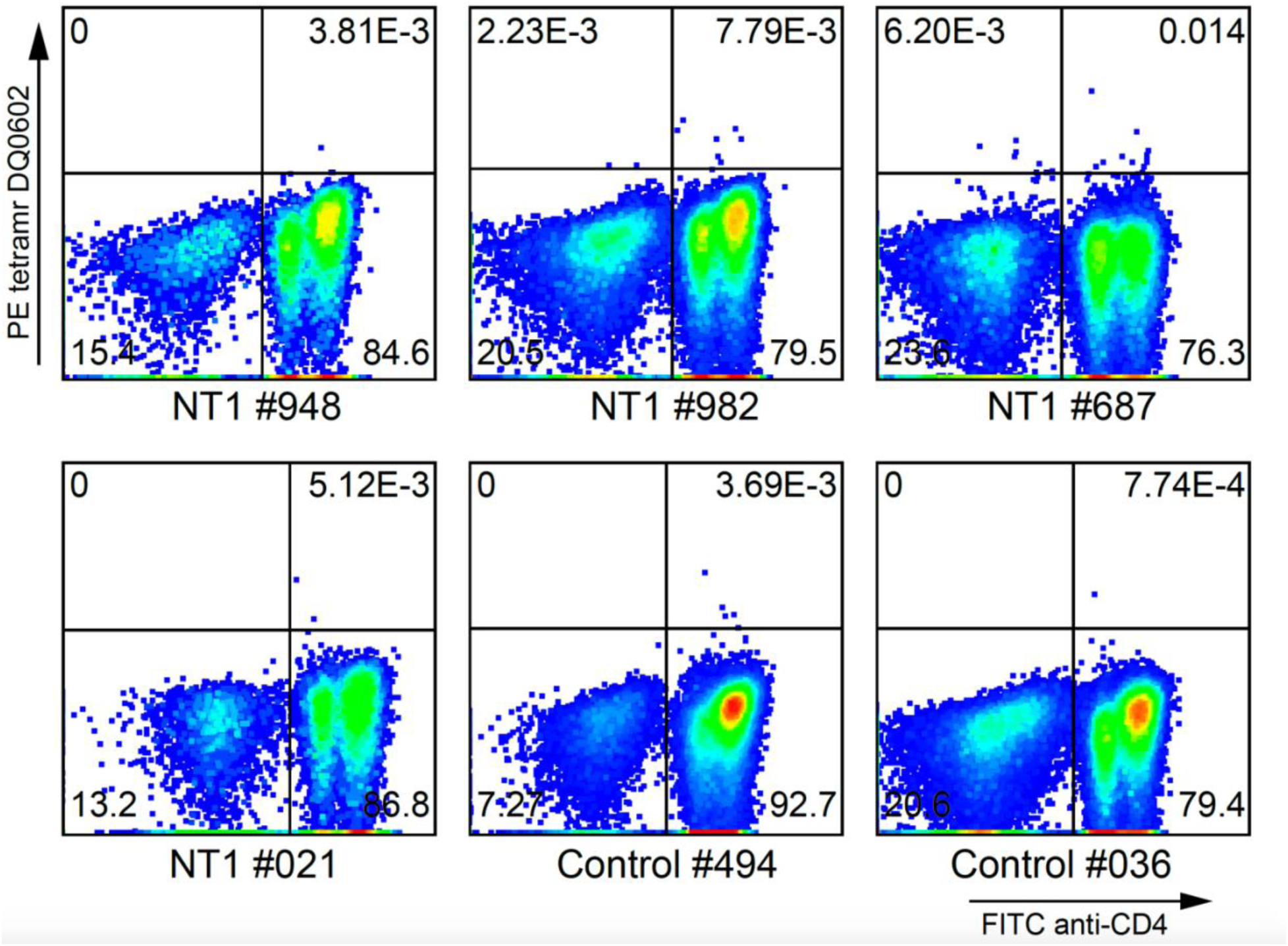
Tetramer staining of T cells recognizing DQ0602 presenting SA<u>PAAASVAPG</u>G, an HCRT epitope known and predicted to bind DQ0602 and predicted to bind DQB1*03:01 in the context of various DQA1 alleles in the same repertoire, although with lower affinity for DQA1*03:03/DQB1*03:01 (see supplementary table 4). Note that very few T cells are found in both controls and NT1, suggesting this reactivity to be rare. Considering these negative results, reactvity to AA<u>PAAASVAPG</u>G bound to DQB1*03:01 associated tetramers was not tested.

